# Work absenteeism across economic activity sectors and its association with COVID-19-like illness prevalence in the Netherlands, 2020-2023

**DOI:** 10.1101/2025.05.26.25328340

**Authors:** Hester Korthals Altes, Jan van de Kassteele, Bram Wisse, Maria Xiridou, Albert Jan van Hoek, Jacco Wallinga

**Author notes:** Corresponding author: Hester Korthals Altes.

## Abstract

The monitoring of work absenteeism can inform pandemic decision making, besides the surveillance of disease end-points like mortality and intensive care bed occupancy. For instance, high disease prevalence accompanied by elevated levels of absenteeism in the healthcare sector will increase the strain on the health care system, and may necessitate adaptation of the control measures. This highlights the need to assess the association between COVID-19 disease prevalence and absenteeism in relevant economic sectors. We initiated the comprehensive monitoring and analysis of work absenteeism and developed an autoregressive time series model which combined COVID-19 prevalence as measured through syndromic surveillance, with absenteeism across various economic activity sectors in the Netherlands. The analysis was updated regularly and shared with policy makers. Overall, prevalence of COVID-19-like illnesses was the most important contributor to variation in absenteeism over the period November 2020-May 2023, with absenteeism rates varying markedly between activity sectors. Of the sectors well-covered by the absenteeism database, the Education and Logistics sectors showed the greatest contribution of a seasonal pattern independent of COVID-19 to absenteeism.

## Introduction

Absenteeism changed over the course of the COVID-19 pandemic. In the Netherlands, the primary focus during the initial stages of the pandemic was to keep up the health care workforce, where the pressure was highest (Mehta et al., 2021; Nehme et al., 2023). In later stages of the pandemic, absenteeism - whether due to actual illness, isolation or quarantine rules-also led to personnel problems in sectors other than healthcare. Disruptions occurred in (transport) services (Zwolsman, 2022), a lack of personnel was seen in the hospitality industry -in addition to problems due to unskilled workers leaving the sector (Notten and Hooijmaaijers, 2021)-as well as in the education sector. The relevance of absenteeism was illustrated by the inclusion of absenteeism as an outcome measured in a number of COVID-19 vaccination studies (Maltezou et al., 2023; Maltezou et al., 2021). The study of COVID-19 incidences in specific economic activity sectors is useful, on a short timescale, to evaluate the need for non-pharmaceutical interventions (NPI’s) in all sectors or only in those most at risk (Verbeeck et al., 2021). On a longer time scale, this may allow the comparison of vaccination effects and other measures (Stijven et al., 2023). In the current study, we are interested in the economic impact of COVID-19 like illness prevalence, and therefore use sector-specific absenteeism as an outcome measure.

Time series of work absenteeism have been used to flag unusual upsurges in absences to signal an infectious disease outbreak (Groenewold et al., 2021), but only a few studies have investigated the association between incidence of COVID-19 or SARS-CoV-2 infection. In the Netherlands, a study at the global level, and a special focus on the education and healthcare sector, found a positive correlation between absenteeism and SARS-CoV-2 notifications (Keet et al., 2024). In Canada, a city-level study found that absenteeism in the largest employer correlated well with wastewater SARS-CoV-2 (Acosta et al., 2023). None of these studies comprehensively analysed the relationship between prevalence of COVID-19 illness or SARS-CoV-2 notifications and absenteeism by economic activity sector.

We combined the weekly reports of absenteeism, overall and across economic activity with weekly prevalence of COVID-19-like illness as inferred from syndromic surveillance. Using a time series regression model of absenteeism with COVID-19-like illness as an explaining variable, we visualised and quantified the relationship between weekly absenteeism by economic activity sector and weekly COVID-19 illness prevalence. This approach was then used to interpret the variation in absenteeism over time, in relation to the variation in COVID-19-like illness.

## Methods

### General approach

We analyse the relationship between the weekly absenteeism frequency, and weekly COVID-19-like illness prevalence, as derived from the Dutch syndromic surveillance *Infectieradar* (www.infectieradar.nl), from the week starting on Monday November 2^nd^, 2020 to the week ending on Sunday May 28th, 2023. We used syndromic surveillance data rather than testing data because testing requirements changed during the study period, and trends in numbers of test-confirmed cases may not reflect actual trends in incidence of COVID-19 illness. For example, a positive self-test was not necessarily to be confirmed by a test at the Municipal Health Services (GGD) after April 11^th^ 2022 (National Institute for Public Health and the Environment, 2023a). We could not include the first wave in the analysis as the syndromic surveillance was not fully operational at that time.

We started producing these analyses when COVID-19 measures were being relaxed, early 2022, while the Omicron variant was present. It was done in a real-time manner as the data was analysed on the Mondays after the last week included in the study, and could be shared with policy-makers on Tuesdays. Results, each time based on data supplemented with two weeks or a month ‘input, were communicated respectively biweekly in early 2022, and later monthly.

### Work absenteeism data

Work absenteeism, defined as weekly notifications of absences due to sick-leave, overall and by economic activity sector (see overview of sectors in Table 1), was made available by HumanTotalCare (HTC), consisting of ArboNed and HumanCapitalCare. This nationwide Dutch occupational health service assists employees and employers by registering amongst others absenteeism, i.e. when an employee is absent for one or more days due to illness. HTC covers approximately 11% of the total Dutch working population as defined by Statistics Netherlands (Statistics Netherlands, 2024a). The coverage per sector was derived from the same database (Statistics Netherlands, 2024a), as well as the age-distribution per sector (Statistics Netherlands, 2024b). Here we used the absenteeism reporting frequency, denoted by *y*(*t*), for the given week *t* per employee. It is measured as the number of new absence reports in that week divided by the total number of employees (absent as well as not absent) in that same week; it is rescaled to a frequency per employee per year. It is an incidence measure of absenteeism.

**Table 1:** Overall percentage contribution of each component to work absenteeism by economic activity sectors included in the study. Coverage of the sectors by the HumanTotalCare data is usually between 5 and 20%; exceptions are indicated with * (<5% coverage) and ? (>20% coverage).

| Economic activity sector | Short name in Figure 2 | Overall percentage contribution of each component to variation in absenteeism |  |  |  |  |
| --- | --- | --- | --- | --- | --- | --- |
|  |  | COVID-19 like illness | Seasonal effect | Long-term trend | Autoregressive effect | Residual |
| A - Agriculture, forestry and fishery * | Primary sector | 33.3 | 25.5 | 17.7 | 0.1 | 23.3 |
| B - Mineral extraction * | Mining | 19.3 | 6.8 | 10.8 | 5.4 | 57.7 |
| C - Industry | Industry | 54.7 | 11.5 | 17.3 | 2.7 | 13.8 |
| D - Production and distribution of and trade in electricity, natural gas, steam and cooled air * | Energy | 29.5 | 11.2 | 10.8 | 3.9 | 44.6 |
| E - Water collection and distribution, waste and sewage treatment ¶ | (Waste) water mgmt | 39.4 | 18.5 | 22.3 | 4.9 | 15 |
| F - Construction | Construction | 34 | 16.3 | 28.6 | 7.7 | 13.4 |
| G - Wholesale and retail; car repair | Retail and repair | 56.1 | 14.4 | 14.4 | 5.9 | 9.2 |
| H - Transport and storage | Logistics | 27.8 | 27.9 | 20.2 | 12.2 | 11.9 |
| I - Lodging, meal and drink supply | Hospitality | 45 | 4.4 | 22.4 | 7.2 | 20.9 |
| J - Information and communication | ICT | 61.1 | 8.1 | 18.7 | 2.1 | 10.1 |
| K - Financial institutions ¶ | Finance | 61.6 | 12.3 | 13.5 | 2.9 | 9.7 |
| L - Rental and trade in real estate | Real estate | 63.1 | 11.7 | 10.8 | 1.3 | 13 |
| M - Advice, research and other specialistic services | Consultancy | 60.5 | 11.5 | 14.4 | 3.8 | 9.8 |
| N - Lease of movable property and other services | Rental services | 48.3 | 15.7 | 21.6 | 3.6 | 10.9 |
| O - Public administration, services, and mandatory social insurances * | Public services | 54.2 | 16.5 | 16.3 | 2.1 | 10.9 |
| P - Education | Education | 21.8 | 22.9 | 26.6 | 8.8 | 19.9 |
| Q - Healthcare and welfare | Healthcare | 60.7 | 18.4 | 9.4 | 4.5 | 7 |
| R - Culture, sports and recreation | Leisure | 51.8 | 0 | 24.7 | 6.7 | 16.7 |
| S - Other services | Other services | 53.6 | 10.6 | 14.7 | 3.2 | 17.9 |
| Total |  | 55 | 14.2 | 17.1 | 5 | 8.7 |

The reporting frequency of absenteeism per person per year for week *t* and for sector *j* is given by *y*(*t, j*) = 52.18 *z*(*t, j*)/*N*(*t, j*), where *z*(*t, j*) is the number of new sick-leave absence reports in week *t* for sector *j* and *N*(*t, j*) is the average number of employees from sector *j* covered by HTC in week *t*. The number 52.18 is the average number of weeks in one year (365.25/7). The reporting frequency over all sectors is computed as *y*(*t*)_*total*_ = 52.18 Σ_*j*_ *z*(*t, j*)/ Σ_*j*_ *N*(*t, j*).

### Syndromic surveillance

The *Infectieradar* syndromic surveillance is a web-based participatory system that was set up in the Netherlands mid-March 2020, and fully operational from November 2020 onwards. It collects data on symptom occurrence, SARS-CoV-2 testing, vaccination against influenza virus and COVID-19, absenteeism, etc. through weekly surveys filled out by the respondents (Bulsink et al., 2020; McDonald et al., 2021a; McDonald et al., 2021b). The data used in the current study is a weekly prevalence measure. It is defined as the weekly average of the percentage of participants reporting COVID-19-like symptoms, per reporting date of a weekly questionnaire. COVID-19-like symptoms were defined as one or more of a set of 19 symptoms (fever, chills, runny or blocked nose, sneezing, sore throat, cough, dyspnoea (shortness of breath), headache, muscle/joint pain, chest pain, malaise, loss of appetite, coloured phlegm, watery or bloodshot eyes, nausea, vomiting, diarrhoea, stomachache, loss of sense of smell/taste, other) within the 7-day period prior to reporting. The data is available online (National Institute for Public Health and the Environment, 2023b).

Participants are allowed to join and leave *Infectieradar* at any time, so the number of participants varies: within the study period, for example, as of June 2022, *Infectieradar* counted roughly 9,000 active participants (McDonald et al., 2024). The age and gender distribution of current and past participants is also indicated on the dashboard (National Institute for Public Health and the Environment, 2023c): overall, women and age categories 50-80 years are overrepresented. Well-represented is the age category 40-50 years; underrepresented are age-categories under 20 and over 80 years. For a more comprehensive description of the syndromic surveillance, see McDonald et al. (2024).

Figure 1 shows the time series of the weekly average percentage of participants reporting COVID-19-like symptoms over the study period -from the start of Infectieradar in November 2020, until the COVID-19 meetings reporting to the ministry were suspended in June 2023. On three occasions spanning 4 weeks, the underlying daily percentage of participants reporting COVID-19-like symptoms was missing (8 days in October 2021, 2 days in June and 2 days in July 2022) due to technical malfunctions of the survey. These missing daily values were imputed by linear interpolation, before aggregating into weekly values.

**Figure 1.**
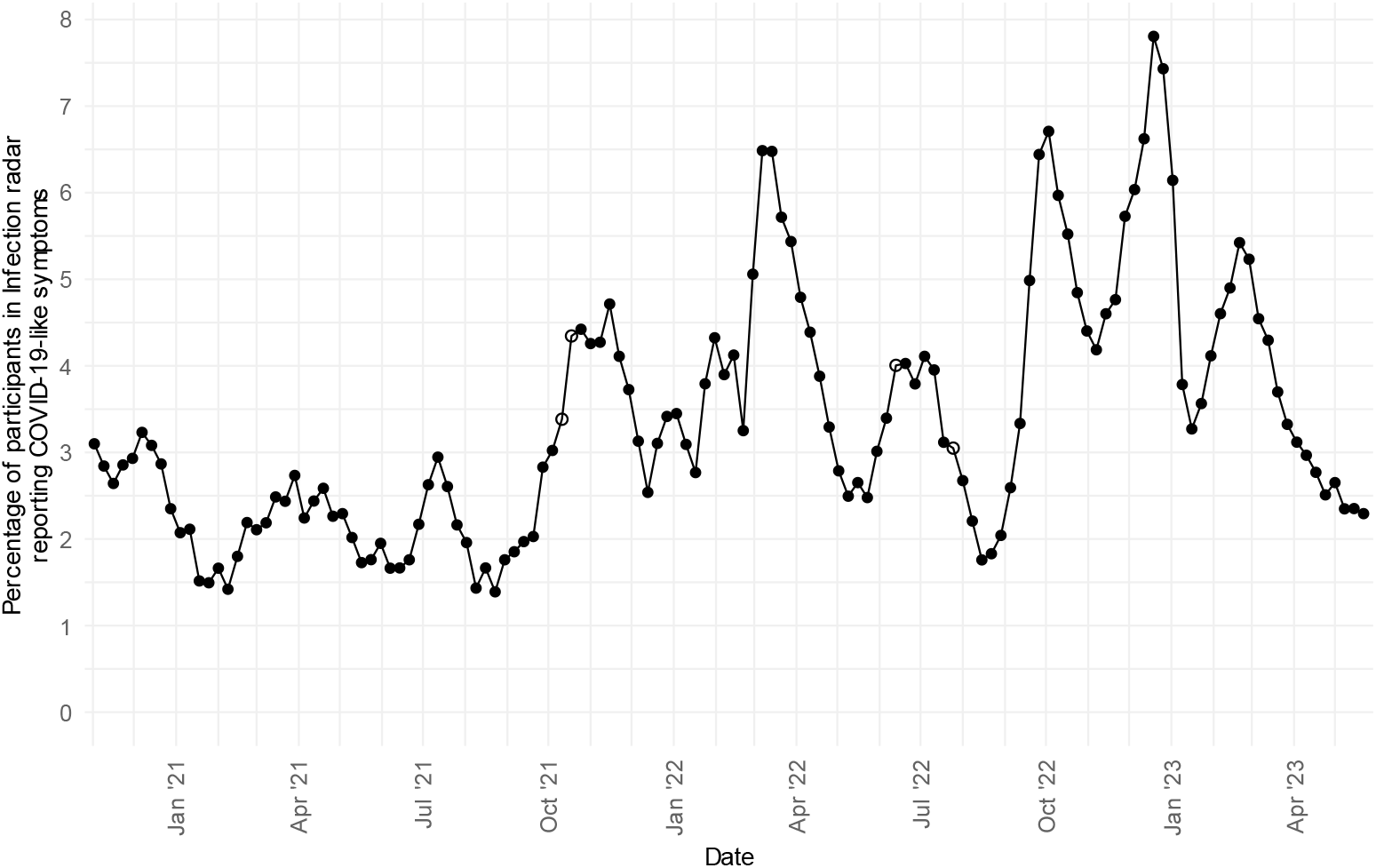
Weekly average percentage of participants reporting COVID-19-like symptoms in Infectieradar in the Netherlands from November 2, 2020 – May 28, 2023. The four open dots represent weeks where the percentage was based on imputed values. Markers are plotted on the first day of the week.

### Statistical model

To analyse the association between the weekly absenteeism reporting frequency in each economic activity sector (and in the total of all sectors), with the percentage *Infectieradar* participants reporting COVID-19-like symptoms in that same week, we used an ARX(*p*) time series regression model (Shumway and Stoffer, 2006). This is an autoregressive time series model of order *p*, with explanatory variables.

It can be written as:

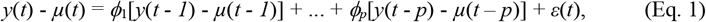

where *µ*(*t*) is a time-varying mean depending on the explanatory variables, *ϕ*_*i*_ are the autoregressive parameters for lag *i* = 1, …, *p*, and *ε*(*t*) is a zero-mean Gaussian white noise error with constant variance *ς*_2_.

The time-varying mean *µ*(*t*) can be written as the sum of: 1) a long-term time trend, also including a Christmas holiday effect, 2) a seasonal effect, and 3) the effect of the weekly percentage *Infectieradar* participants reporting COVID-19-like symptoms:

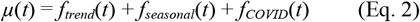

The long-term time trend *f*_*trend*_(*t*) was described as a constant, a straight line, or a natural cubic spline with two degrees of freedom. Additionally, the trend term always included Christmas holidays (two school holiday weeks including Christmas day and New Years day), coded as a binary variable. The seasonal effect *f*_*seasonal*_(*t*) was described by a sum of sine and/or cosine functions with a period of one year (52.18 weeks), and where needed with sine and/or cosine functions with a period of six months. A seasonal effect was not always included in the model: for some activity sectors, the model fit was better without seasonal component. The relation between the prevalence of *Infectieradar* participants with COVID-19-like symptoms and absenteeism, *f*_*COVID*_(*t*), was described by a linear relation, or a natural cubic spline with two degrees of freedom. This effect was always included in the model. The autoregressive effect could have order *p* set to 1, 2, 3 or 4, in other words, *y*(*t*) - *µ*(*t*) may depend on its previous values up to four weeks back.

Based on these combinations, in total 312 models were considered for each economic activity sector. The best model was chosen based on the lowest Akaike Information Criterion (AIC).

Each best fit model was post-processed for visualisation of the results. First, we calculated the total autoregressive effect *f*_*AR*_(*t*) (the right hand side of Eq. 1, minus *ε*(*t*)), next to the three components *f*_*trend*_(*t*), *f*_*seasonal*_(*t*), and *f*_*COVID*_(*t*). Then, in order to create a stacked plot of these four components, we subtracted the minimum value of each component from the component values itself, making sure the lowest value of each component was always greater than zero, and thus making stacking of the components possible for visualisation. To compensate for these four subtractions, the sum of the minimum values was added to each value of the time trend component. For this reason, the visualised time trend in the plots could be lower than zero. Note that this is purely an artefact of the post-processing step.

To obtain a rough estimate of the proportion of variance explained by each component, we calculated the variance of each component over the full study period, and divided it by the sum of all component variances.

Data handling, model fitting and postprocessing of the results were done in R (R Core Team, 2023). The data and code for the analysis is available online: https://github.com/kassteele/Papers.

## Results

Figure 2 shows the reported absenteeism and the results of the model fits per sector and for the total of all sectors. The blue-shaded areas represent the components as estimated by the time series regression model, contributing to the variation in absenteeism.

**Figure 2.**
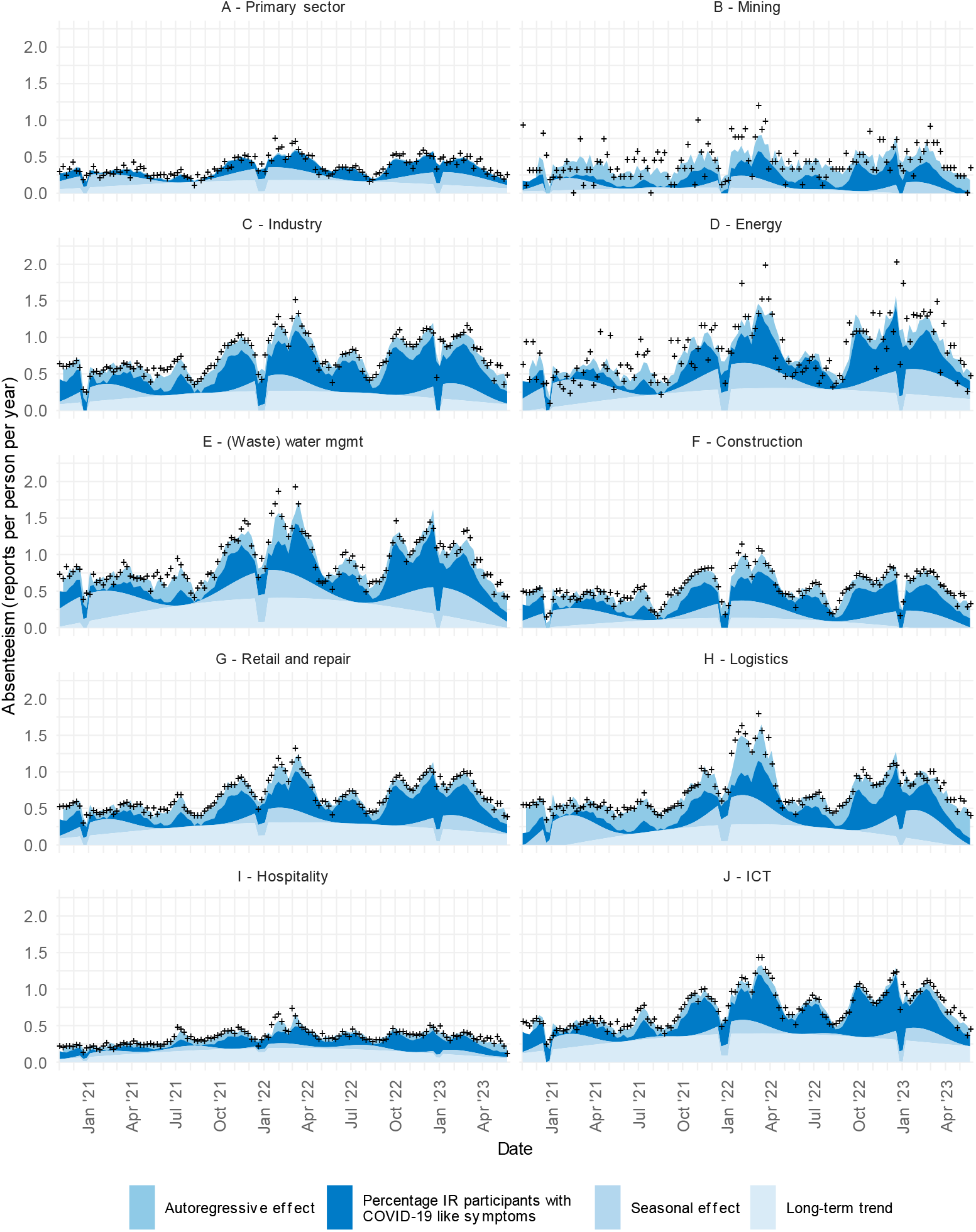
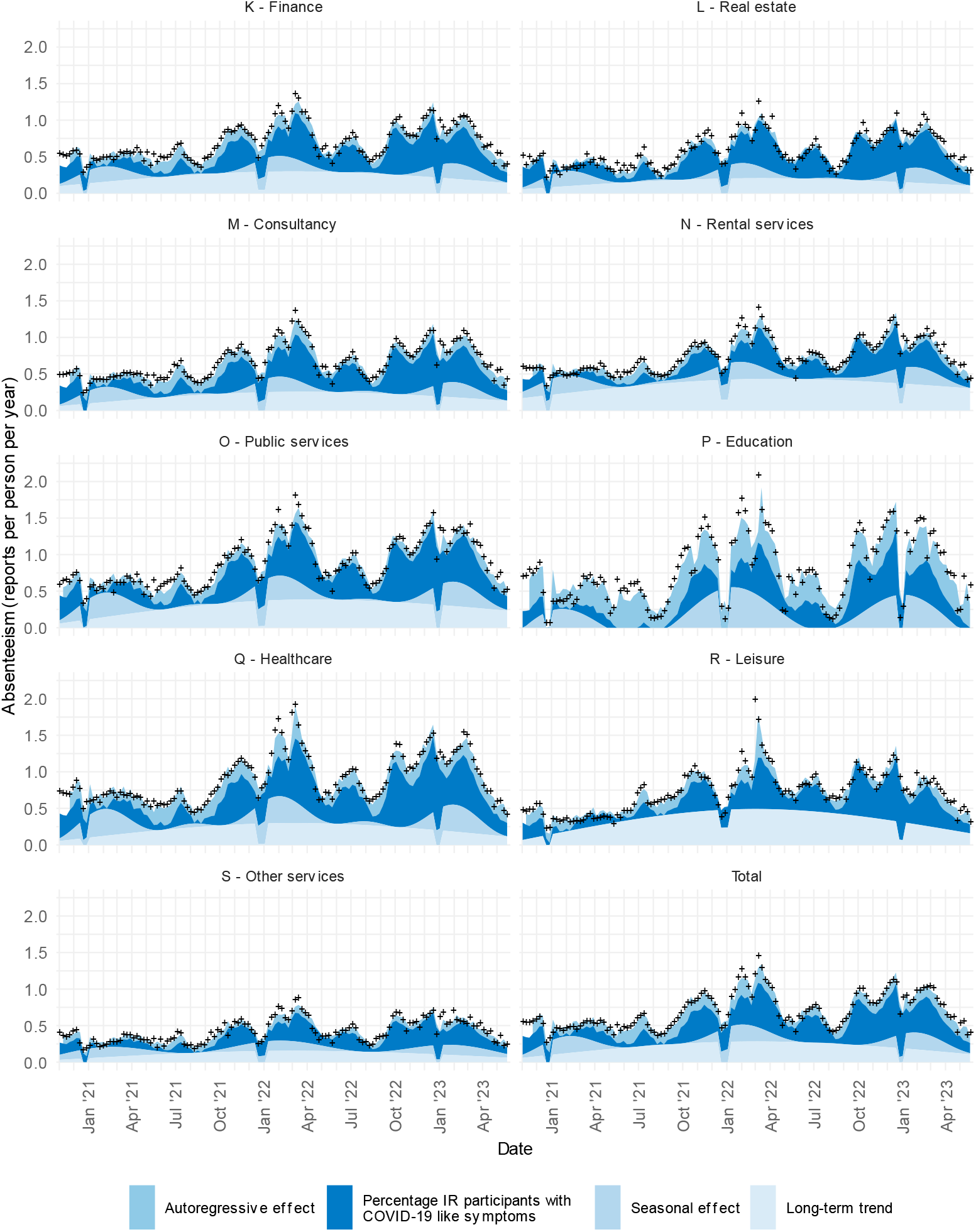
Absenteeism reporting frequency (per employee per year) per economic activity sector in the Netherlands from November 2, 2020 – May 28, 2023. Markers: reported absenteeism (HumanTotalCare data). Shaded areas: proportion of absenteeism explained by the components of the statistical model (Eq. 1 and Eq. 2). The four components are given in the legend.

Overall absenteeism (Figure 2, bottom right panel) shows substantial variation between November 2020 and June 2023, with smaller and larger peaks: the lowest average reporting frequency is at 0.27 reported sick leaves per person per year (Fig. 2 Total: in week starting Dec 21, 2020), and with average notification frequencies peaking at 1.46 reported sick leaves per person per year (first week of March 2022). The long-term trend in absenteeism (lightest shade of blue) increases between November 2020 and April 2022, and then declines until June 2023. The trend is interrupted by dips in the Christmas holidays. On top of this trend, the seasonal pattern in absenteeism is seen (darker shade): three seasonal peaks occur, corresponding with each winter. The darkest shaded area represents the share of absenteeism associated with the percentage respondents that reported COVID-19-like illness. In peak absenteeism periods, COVID-19-like illness explains most variation in absenteeism. The uppermost shaded area represents the autoregressive component. This is the variation explained by absenteeism reported in the previous week(s), and which could not be explained by the other three components.

The dynamics of absenteeism, *i*.*e*. levels of reporting and patterns over time, differ between economic activity sectors (Figure 2). Four examples of sectors with high absenteeism are (Waste) Water management (mgmt), Public services, Education, and Healthcare, showing peaks in the first week of March 2022 at frequencies around 1.5-2 reported sick leaves per person per year (Figs. 2E, 2O, 2P, 2Q; peaks at respectively 1.93, 1.81, 2.08, 1.92 reported sick leaves per person per year). In contrast, absenteeism stays below 1 reported sick leave per person per year in the Primary sector, Hospitality sector, and Other services (Figs. 2A, 2I and 2S; peaks at respectively 0.74, 0.74, and 0.88 reported sick leaves per person per year, between January and March 2022). The long-term time trend in absenteeism is similar across sectors, increasing in the period between November 2020 until the first half of 2022, and (slightly) decreasing up to June 2023. There generally is a yearly seasonal trend in absenteeism, with a low-season between May and September, and a high season between January and March. This seasonal trend is stronger in some sectors, like Education and-Healthcare, than in other sectors, like ICT; it is almost absent in the Hospitality sector, and absent in the Leisure sector. Some economic activity sectors have a comparatively larger autoregressive component for example in the sectors Construction, Logistics, and Education. Absenteeism peaks at a much higher frequency in many sectors in 2022 than in 2023 (for example Figs. 2E, 2H, 2Q, 2R).

In 11 out of the 19 sectors represented, variation in prevalence of COVID-19 like-illness contributes more than 50% to the variation in absenteeism, averaged over the whole study period (Table 1). This is also the case for the variation in absenteeism in all sectors combined (Table 1). The sectors with greatest association -over 60%-between prevalence of COVID-19-like illness and absenteeism, are Real Estate, Finance, ICT, Consultancy and Healthcare (respectively 63.1%, 61.6%, 61.1%, 60.5% and 60.7%, see Table 1).

## Discussion

To our knowledge, this is the first study quantifying the association between work absenteeism per economic activity sector and COVID-19-like illness prevalence. It applies the methods that we used for real-time analyses carried out for policy-making briefs starting in 2022. We observed a positive association, in all sectors, between absenteeism and COVID-19-like illness, after accounting for different seasonal and long-term trends in absenteeism. The autoregressive component differed between sectors, reflecting the differences in impact of other factors, including other infectious diseases and the COVID-19 cases not captured by *Infectieradar*. This could explain in some instances why the autoregressive component is greater during the COVID-19 peaks than in the troughs.

Overall, during the period of containment of the SARS-CoV-2 infections (2020-2021), absenteeism was lower than in the following two years (2022-2023). The relaxation of measures in 2022 associated with the Omicron variant led to very high peaks in COVID-19-like illness, which in turn gave rise to high absenteeism levels. The peak absenteeism reporting frequencies observed in 2022 and 2023 were comparable to those observed during a very severe influenza season, such as in early 2018 (ArboNed, 2022). However, the duration of the elevated absenteeism period was much longer in 2022-2023 than in 2018.

Our general observation is that absenteeism is associated with COVID-19-like illness, but the analysis does not reveal whether the association changed over time. An extension of this work would be to test whether COVID-19-like illness contributed more to absenteeism in some phases of the study period than in others. The auto-regressive model should then be adapted to allow for a time-interaction term or time-variant parameters in different phases (for example variant-periods as in Keet et al. (2024)). A city-level study in Canada that related absenteeism with wastewater surveillance trends observed a stronger relation between COVID-19-related absenteeism and SARS-CoV-2 RNA concentrations in wastewater during the Omicron peak than in the prior peaks (Acosta et al., 2023).

We have identified a positive association at the population level between prevalence of COVID-19 like illness and absenteeism overall and per economic activity sector. The data do not allow to establish a similar association at a finer level. We could not split the prevalence of COVID-19-like illness by economic activity sector, as this information was not available. We used the data from all syndromic surveillance participants, regardless of age; this means that we related absenteeism, which only applies to working individuals, to COVID-19-like illness reported in the Infectieradar cohort. *Infectieradar* has an overrepresentation of people aged 40-70 years, while the younger and elderly are underrepresented (National Institute for Public Health and the Environment, 2023c). Since the time series for the percentage COVID-19-symptomatics in 25-39 year-olds versus 60-69 year-olds (National Institute for Public Health and the Environment, 2023c) follow a similar temporal pattern, with the younger age group only occasionally showing a peak or trough a week before it occurs in the older age group, we suspect that the association will also hold specifically for the working age-groups. Regardless, our main conclusion of a strong association between absenteeism and COVID-19 prevalence at a population level is not affected.

A Dutch study showed how the measure from syndromic surveillance we used here can be attributed to Influenza and SARS-CoV2 over time (McDonald et al., 2024). It appears that over 2022 for example, most of COVID-19 like illness was attributable to SARS-CoV2 infections until week 10; then Influenza became the major contributor to COVID-19-like illness. From week 17 onwards, yet other respiratory pathogens became more important. The scope of our study is to investigate the relationship between absenteeism across economic activity sectors and COVID-19-like illness. That the latter may include disease caused by other respiratory pathogens is less problematic for our purposes, than trying to use alternative, possibly more specific measures for COVID-19-like illness. This is because the background of our study question is to follow, as the pandemic was unfolding, to what extent the use of NPI’s, and their relaxation -which also influences circulation of other respiratory pathogens-contributed to upsurges in absenteeism through increases in COVID-19-like illness. Other measures come with other methodological challenges: wastewater surveillance gives estimates for SARS-CoV2 infections, but gives no indication of how many people had symptoms and therefore took sick leave. Hospitalization data, on the other extreme, has the caveat that severity of disease/hospitalization rates differ by SARS-CoV2 variant and by vaccination coverage. This means that one might observe a similar incidence of hospitalized COVID-19 cases for different time periods, but quite different corresponding incidence of mild COVID-19 disease, which in turn would be associated with different levels of work absenteeism. In short, we focused on a small part of the causal chain of events, i.e. that between the prevalence of COVID-19-like illness and absenteeism.

Finally, the nature of absenteeism, such as the average duration of and the reason for sick leave, might have changed over the course of the pandemic. A Dutch study using the same data source for absenteeism as we did, examined the employee-related and SARS-CoV-2 variant determinants of the return-to-work rate: a faster return-to-work rate was observed in the periods with milder variants dominating (Aben et al., 2023). It might therefore be interesting to also look both at the average net percentage absenteeism (a measure taking into account sick leave duration), that has increased steadily from 2015-2022 (Statistics Netherlands, 2023), as well as the distribution of sick leave duration over the course of the pandemic. In combination with the previous suggestion of analysing the absenteeism by variant period, this could shed light on the possible impact of differential severity of disease for example, on absenteeism.

This work shows that the dynamics and levels of absenteeism differ across economic activity sectors, but a consistent and strong association between absenteeism and COVID-19-like illness prevalence is observed. This association could allow for short-term projections of work absenteeism based on COVID-19 prevalence. Bringing together these epidemiological and socio-economic data streams can help to inform public health policy makers about the extent to which absenteeism could be due to COVID-19.

## Data Availability

The source data on COVID-19 syndromic surveillance are available on: https://dashboard.infectieradar.nl/ 
The absenteeism data is available upon request from the co-author at HTC (Bram Wisse)

## References

Aben, B., Kok, R.N., de Wind, A., 2023. Return-to-work rates and predictors of absence duration after COVID-19 over the course of the pandemic. Scand J Work Environ Health 49, 182–192.

Acosta, N., Dai, X., Bautista, M.A., Waddell, B.J., Lee, J., Du, K., McCalder, J., Pradhan, P., Papparis, C., Lu, X., Chekouo, T., Krusina, A., Southern, D., Williamson, T., Clark, R.G., Patterson, R.A., Westlund, P., Meddings, J., Ruecker, N., Lammiman, C., Duerr, C., Achari, G., Hrudey, S.E., Lee, B.E., Pang, X., Frankowski, K., Hubert, C.R.J., Parkins, M.D., 2023. Wastewater-based surveillance can be used to model COVID-19-associated workforce absenteeism. Sci Total Environ 900, 165172.

ArboNed, 2022. Recordaantal verzuimmeldingen door griepgolf en corona. https://www.arboned.nl/nieuws/persbericht-recordaantal-verzuimmeldingen-door-griepgolf-en-corona (accessed 01-02-2024)

Bulsink, K., Petit, F., Dalhuisen, T., Nijman, S., Jenniskens, L., Smit, T., Berry, D., Wong, D., van Weert, Y., van Hoek, A.J., 2020. Infectieradar, https://data.rivm.nl/meta/srv/eng/catalog.search#/metadata/7bd9dfbf-6b30-47a4-a279-fee4a0964400 (accessed 05-06-2023).

Groenewold, M.R., Free, H., Mobley, A., 2021. Using Workplace Absences to Measure How COVID-19 Affects America’s Workers. CDC, NIOSH Science Blog. https://blogs.cdc.gov/niosh-science-blog/2021/03/12/covid-absences/ (accessed 11-9-2023)

Keet, M.G., Boudewijns, B., Jongenotter, F., van Iersel, S., van Werkhoven, C.H., van Gageldonk-Lafeber, R.B., Wisse, B.W., van Asten, L., 2024. Association between work sick-leave absenteeism and SARS-CoV-2 notifications in the Netherlands during the COVID-19 epidemic. Eur J Public Health. doi: 10.1093/eurpub/ckae051.

Maltezou, H.C., Basoulis, D., Bonelis, K., Gamaletsou, M.N., Giannouchos, T.V., Karantoni, E., Karapanou, Α., Kounouklas, K., Livanou, M.E., Zotou, M., Rapti, V., Stamou, P., Loulakis, D., Souliotis, K., Chini, M., Panagopoulos, P., Poulakou, G., Syrigos, K.N., Hatzigeorgiou, D., Sipsas, N.V., 2023. Effectiveness of full (booster) COVID-19 vaccination against severe outcomes and work absenteeism in hospitalized patients with COVID-19 during the Delta and Omicron waves in Greece. Vaccine 41, 2343–2348.

Maltezou, H.C., Panagopoulos, P., Sourri, F., Giannouchos, T.V., Raftopoulos, V., Gamaletsou, M.N., Karapanou, A., Koukou, D.M., Koutsidou, A., Peskelidou, E., Papanastasiou, K., Souliotis, K., Lourida, A., Sipsas, N.V., Hatzigeorgiou, D., 2021. COVID-19 vaccination significantly reduces morbidity and absenteeism among healthcare personnel: A prospective multicenter study. Vaccine 39, 7021–7027.

McDonald, S.A., Jan van Hoek, A., Paolotti, D., Hooiveld, M., Meijer, A., de Lange, M., Infectieradar team, van Gageldonk-Lafeber, A., Wallinga, J., 2024. A statistical modelling approach for determining the cause of reported respiratory syndromes from internet-based participatory surveillance when influenza virus and SARS-CoV-2 are co-circulating. PLOS Digit Health 3, e0000655.

McDonald, S.A., Soetens, L.C., Schipper, C.A., Friesema, I., van den Wijngaard, C.C., Teirlinck, A., Neppelenbroek, N., van den Hof, S., Wallinga, J., van Hoek, A.J., 2021a. Testing behaviour and positivity for SARS-CoV-2 infection: insights from web-based participatory surveillance in the Netherlands. BMJ Open 11, e056077.

McDonald, S.A., van den Wijngaard, C.C., Wielders, C.C.H., Friesema, I.H.M., Soetens, L., Paolotti, D., van den Hof, S., van Hoek, A.J., 2021b. Risk factors associated with the incidence of self-reported COVID-19-like illness: data from a web-based syndromic surveillance system in the Netherlands. Epidemiology and Infection 149, e129.

Mehta, S., Machado, F., Kwizera, A., Papazian, L., Moss, M., Azoulay, É., Herridge, M., 2021. COVID-19: a heavy toll on health-care workers. The Lancet Respiratory Medicine 9, 226–228.

National Institute for Public Health and the Environment (RIVM), 2023a. Coronavirus Timeline. https://www.rijksoverheid.nl/onderwerpen/coronavirus-tijdlijn (accessed 05-06-2023)

National Institute for Public Health and the Environment (RIVM), 2023b. Covid-19 Percentage deelnemers met COVID-19-achtige klachten in Infectieradar per rapportagedatum, 06-06-2023 https://data.rivm.nl/meta/srv/eng/catalog.search#/metadata/7bd9dfbf-6b30-47a4-a279-fee4a0964400 (accessed 05-06-2023)

National Institute for Public Health and the Environment (RIVM), 2023c. Dashboard Infectieradar; Verdeling leeftijd en geslacht van Infectieradar deelnemers. https://dashboard.infectieradar.nl/ (accessed 05-06-2023)

Nehme, M., Vieux, L., Kaiser, L., Chappuis, F., Chenaud, C., Braillard, O., Courvoisier, D.S., Reny, J.-L., Assal, F., Bondolfi, G., Graf, C., Zekry, D., Stringhini, S., Spechbach, H., Jacquerioz, F., Salamun, J., Lador, F., Guerreiro, I., Coen, M., Agoritsas, T., Benzakour, L., Genevay, S., Lauper, K., Meyer, P., Poku, N.K., Landis, B.N., Grira, M., Allali, G., Vetter, P., Guessous, I., HealthCo Study, T., 2023. The longitudinal study of subjective wellbeing and absenteeism of healthcare workers considering post-COVID condition and the COVID-19 pandemic toll. Scientific Reports 13, 10759.

Notten, F., Hooijmaaijers, S., 2021. Corona en de horeca. Statistics Netherlands (CBS). https://www.cbs.nl/nl-nl/longread/de-nederlandse-economie/2021/corona-en-de-horeca?onepage=true

Shumway, R.H., Stoffer, D.S., 2006. Time Series Analysis and Its Applications With R Examples in R, 2nd ed. Springer, New York.

R Core Team, 2023. R: A Language and Environment for Statistical Computing. R Foundation for Statistical Computing, Vienna, Austria. https://www.R-project.org/

Stijven, F., Verbeeck, J., Molenberghs, G., 2023. Comparing COVID-19 incidences longitudinally per economic sector against the background of preventive measures and vaccination. Biometrics 79, 2516–2524.

Verbeeck, J., Vandersmissen, G., Peeters, J., Klamer, S., Hancart, S., Lernout, T., Dewatripont, M., Godderis, L., Molenberghs, G., 2021. Confirmed COVID-19 Cases per Economic Activity during Autumn Wave in Belgium. Int J Environ Res Public Health 18.

Zwolsman, N., 2022 Minder bussen, metro’s en treinen door hoog ziekteverzuim, De Nieuwe Nederlandse Courant (NRC), 28 Jan. 2022. https://www.cbs.nl/nl-nl/longread/de-nederlandse-economie/2021/corona-en-de-horeca?onepage=true

